# The maternal and perinatal health data system in the Kilimanjaro Region, Tanzania: a landscape review including a qualitative study

**DOI:** 10.1101/2024.09.24.24314329

**Authors:** Modesta Mitao, Francis M. Pima, Lucy Munishi, Mary Cronin, Gaudensia A. Olomi, Allen Lyimo, Allen Senkoro, Gustarv Nkya, Jackline Somi, Daire Buckely, Yudathadeus Kyara, Anne Shuma, Fatina Rashid, Bariki Mchome, Michael Johnson Mahande, Jairy Khanga, Jane Hirst, Christopher Redman, Simon Woodworth, Blandina T. Mmbaga, Ali S Khashan

**Author notes:** These two authors contributed equally and are joint first authors (Modesta Mitao, Francis M. Pima). Died on the 12th of August 2024 (Christopher Redman). Corresponding author: MM.

## Abstract

**Background:** The World Health Organization (WHO) advocates the use of digital health interventions to strengthen health systems globally. This landscape review evaluated how data related to maternal and neonatal health is recorded and utilized for clinical and policy decision-making in the Kilimanjaro Region of Tanzania.

**Method:** This landscape review employed a dual method approach, initially quantifying data recording practices of each clinic and then using qualitative interviews to explore how data is currently captured and utilized. We conducted interviewed with seven Nurse-Midwives, eight Lead Obstetricians at district hospitals, seven District Medical Officers, and seven District Reproductive and Child Health Coordinators. Qualitative data analysis was performed using NVivo software.

**Results:** In the region’s seven districts, there are 489 healthcare facilities offering Reproductive and Child Health Services, delivery, infant and child care. The facilities comprise 22 hospitals, 62 health centers, 353 dispensaries, and 52 clinics. In 2021, there were 45,343 births with a maternal mortality rate of 146 per 100,000 live births, a neonatal mortality rate of 10.1 per 1,000 live births, and a stillbirth rate of 4 per 1,000 births. Additionally, 20.3% of births were delivered via C-section.

Although there was a notable decline in maternal and neonatal mortality rates (from 112 to 51 vs 9.5 to 3.7 deaths per 100,000 live births) from 2020 to 2023, the 2021 data revealed unexpectedly high maternal mortality rates. The qualitative study highlighted issues with the data recording system, which is complex and time-consuming, involving multiple paper-based and digital records. Challenges include data volume, loss, and retrieval issues, affecting clinical decision-making. This data is crucial for identifying resource needs, monitoring health trends like low antenatal care uptake, and advocating for changes in resources, guidelines, and policies at regional and national levels.

**Conclusion:** Quantitative data indicates a significant rise in maternal and neonatal mortality rates in the Kilimanjaro region during 2020-2021, followed by a decrease in 2022-2023. Additionally, early antenatal care (ANC) uptake remains below 50%. The qualitative study suggests that paper records are time-consuming and prone to incomplete data making it difficult to use due to their storage approach and the risk of damage to the physical books.

## Introduction

Perinatal health is defined as the health of women and babies before, during, and after six weeks after birth. The WHO recommends the use of digital health globally and within countries to improve healthcare services. Towards attaining universal health coverage and the health-related Sustainable Development Goals, perinatal health information in electronic systems need to be emphasized to reduce perinatal deaths (WHO, 2015; WHO, 2021).

The global maternal mortality is estimated to be 223 deaths per 100,000 births (WHO, 2021), while in Sub-Saharan Africa the rate was estimated to be 536 deaths per 100,000 live births. According to the WHO report from 2021, the maternal mortality rate in Tanzania was 238 deaths per 100,000 live births (WHO, 2021). However, data from the Ministry of Health (MoH) of Tanzania reports a significantly lower maternal mortality rate of 104 deaths per 100,000 live births (Ministry of Health (MoH)Tanzania Mainland *et al*., 2022).

In 2020, the neonatal mortality rate was 27 per 1000 live births in Sub-Saharan Africa (SSA) (WHO, 2020). A substantial variation in neonatal mortality in SSA can be explained by the quality of healthcare and socioeconomic deprivation (Kayode *et al*., 2017). According to the WHO data, Tanzania had 43 newborn deaths per 1000 live births (WHO, 2020). In Tanzania, by 2021 the under-five mortality rate was reported to be 47 per 1000 births (UNICEF, 2020).

In low-income countries, health information including perinatal health records is documented inadequately. This is a major challenge in efforts to improve outcomes in many of these countries (Kumar and Mostafa., 2020). This leads to estimates based on data with inadequate quality, creating challenges for the design, implementation, and evaluation of interventions. Where documentation of good quality data exists, it is paper-based, which is difficult to summarize or analyze (Latifov and Sahay., 2013).

In Tanzania, the Health Information Management System (HMIS) was established in 1993. In 2007, the Ministry of Health (MoH) in Tanzania Mainland resolved to digitalize its health information access and assist routine operations, management, and decision-making. The MoH in Collaboration with the University of Dar-Es-Salaam and the University of Oslo was involved in sketching and integrating HMIS named District Health Information Software 2 (DHIS2). The data are recorded at the individual level at all facilities and then reported to the health system in aggregate forms using the DHIS2 system; though, it is not known how precise this aggregation process reflects the individual-level data.

The major objective of HMIS is to enable everyone involved in the health sector at the health facility, district, regional, and Ministry of Health (MoH) levels to access and use reliable data. This leads to evaluating the implementation of health policies and guidelines for planning and providing quality services of treatment and prevention at affordable cost (MOHSW, 2013).

There is consensus that Electronic Health Record Systems(EHRSs) enable us to capture, analyze and use the recorded data for monitoring population health and providing patient-centered healthcare services (Jawhari *et al*., 2016; Syzdykova *et al*., 2017). We are currently piloting the ULTRA data recording system for maternal and child health data to support a transition from traditional MTUHA paper-based records (Woodworth et al, 2022). This landscape review aimed to 1) describe the data recording system for pregnancy care and births across health facilities in the Kilimanjaro Region; 2) describe the distribution of births and adverse pregnancy outcomes, this will be a foundation for evaluating the newly established of the birth registry, which is intended to reduce maternal and neonatal deaths through improved quality data in Kilimanjaro region; and 3) Explore healthcare staff’s perceptions, experiences, and expectations regarding the current data recording system. The baseline data was used to illustrate the current burden of maternal and neonatal complications which will be further analyzed using the established electronic system.

## Methods

### Study design

This landscape review included both a cross-sectional study and a qualitative study conducted for the period of one year.

### Study setting

This study was conducted in the Kilimanjaro region, which encompasses seven districts: Moshi District (rural), Moshi Municipal (Urban), Siha, Mwanga, Same, Rombo, and Hai. According to the 2022 national census, the region has a population of 1,861,934 and experiences approximately 44,000 births annually. The region is served by a regional referral hospital and a zonal hospital, the latter providing care for Kilimanjaro as well as the broader Northern Tanzanian region, totaling a population of 15 million people.

### Cross-sectional study

#### Study population, data source, and data collection procedures

We described the hierarchy of maternal and neonatal health facilities in the region and the range of services provided to pregnant women and their babies at each level of the health system. We also described the data collected throughout pregnancy and birth in the current Health Management Information System (HMIS). Our review involved analyzing aggregate maternal and neonatal data from reproductive and child health department at the regional office, including routine records from 2020, 2022 and 2023. These data were accessed by reviewing DHIS2 records from regional office’s reproductive and child health department.

The data are recorded using the nationally standardized paper-based MTUHA books and electronic systems in some health facilities. These MTUHA books include comprehensive information on pregnancy, birth, and child data, which are aggregated/compiled monthly and sent to the Regional Medical Office from each health facility. We collected data on births, first and subsequent antenatal care visits, live births, and stillbirths, maternal deaths, and the number of Cesarean sections, providing an overall report. During antenatal visits, gestational age was estimated using Last Normal Menstrual (LNMP) period, fundal height measurements and ultrasound assessments.

Maternal death refers to death of a woman during pregnant or within 42 days of termination of pregnancy, irrespective of the duration and site of the pregnancy or its management but not from unintentional or incidental causes (United Nations: Department of Economic and Social Affairs, 2022). Stillbirth is defined as a baby born with no sign of life after 28 weeks of completed gestation (UNICEF, 2023). These definitions are essential for accurately interpreting and analyzing the data in the MTUHA books (Data from the Kilimanjaro regional office date 13 June 2024).

### Qualitative study

#### Study participants, data collection methods and procedures

As work on the ULTRA project evolved, the team recognized that a qualitative study would significantly enhance understanding of the current, primarily paper-based data system, and its usage. It was decided to gather data from a sample of healthcare staff involved in perinatal data collection, recording, analysis, or utilization. This involved recruiting one participant from each of the following staff groups across the seven districts in Kilimanjaro: Nurse-Midwives (NMs), District Reproductive and Child Health coordinators (DRCHCOs), Lead Obstetricians (LOs), and District Medical Officers (DMOs). Due to the presence of two hospitals in each district, a second lead obstetrician was recruited, resulting in a total sample of 29 participants. Recruitment was facilitated and supported by the Regional Medical Office, with coordination led by the Regional Nurse Officer (RNO) who oversees all health research activities in the region.

Face-to-face, individual semi-structured interviews were selected as the most appropriate data collection method (Ryan *et al*, 2009), following discussions with potential participants it was agreed that the interviews would be carried out a t their workplaces. Appointments were arranged via phone calls, and most interviews took place after working hours except with DMOs.

Informed, written consent was obtained from each participant and interviews were guided by a pre-tested topic guide; the duration the interviews ranged from 15 to 45 minutes, with those involving being shorter due to their demanding schedules.

The interview was conducted by an experienced post-graduate researcher (MPH), was supported by a research assistant with a Bachelor of Arts in Community Development who acted as note taker, the interviews were conducted in Kiswahili and audio recorded. Qualitative data collection took place in November 2022 to March 2023.

### Data analysis

Project documents were translated into a local language and verbal consent was obtained when necessary. Permission for the secondary use of existing data was sought, where applicable. Descriptive analysis was performed, with categorical variables summarized using frequencies and proportions. Mean and confidence intervals were calculated and presented. Figures and tables were used to present the data. Qualitative research transcripts were translated into English and thematic analysis (Braun and Clarke, 2006; Braun and Clarke, 2022), was completed collaboratively by four researchers, with the aid of NVivo 12 software.

### Data availability

Researchers interested in accessing data from the Regional Administrative office should contact the Regional Administrative secretary of the Kilimanjaro region. Due to privacy and ethical considerations, the data are not publicly available and can only be accessed upon reasonable request and ethical considerations, the data are not publicly available and can only be accessed upon reasonable request.

### Ethical considerations

This study involving human participants was conducted following all applicable ethical guidelines and regulations. The detailed protocols and consent documents were drafted and the approval was granted by the College Research Ethics Review Committee of Kilimanjaro Christian Medical University College (approval number 2606) and the Research Ethics Committees in Tanzania (ethical approval number NIMR/HQ/R.8a/Vo1. IX/4075). Project documents were translated to the local language, and verbal consent when needed. Permission for use of secondary data was sought, where applicable. Given that pregnant women and new mothers are considered vulnerable populations, we ensured that their consent was fully informed. All project researchers received training in Good Clinical Practice including specific training on obtaining consent. Explicit consent was sought for the collecting of sensitive personal information (clinical/ health), either in written or verbal form. All data collection, storage, transfer and analysis adhered to the Data Protection Acts 1988 and 2003 concerning the protection of personal data, on the Protection of Privacy of Individuals with regard to Personal Data. The study complied with the Declaration of Helsinki principles. As well as to Data Protection Data Protection Guidelines in Tanzania. Qualitative data was stored securely in a university online data space group account which was accessible only to the qualitative research study team.

## RESULTS

### Cross-sectional

#### Health facilities

In Kilimanjaro region, there are 489 healthcare facilities categorized into hospitals, health centers, dispensaries, and clinics. The majority, comprising 72.2% (353 out of 489), are dispensaries, health facilities 72.2% (353/489) are dispensaries. Ownership of these facilities is distributed among various stakeholders, including government entities, faith-based organizations (FBOs), and private owners. Over half of the healthcare facilities are government-owned, totaling 57.7% (282 out of 489). The distribution of facilities is as follows: 22 hospitals, 62 health centers, 353 dispensaries, and 52 clinics (**Table 1 & Figure 1**).

#### Electronic system

In the region, hospitals use electronic health record systems like e-HIMS and Afya Care for patient registration upon admission. District hospitals utilize e-HIMS to capture patient details such as name, date of birth, age, sex, residence, marital status, phone number and insurance status. However, there is no integration of information for the maternal and child department. Health centers and dispensaries rely solely on a paper-based system for recording data. Additionally, all hospitals at each level record maternal and neonatal information using MTUHA books.

#### Health services

The reproductive and child health services provided include ANC services, CECAP (Cervical cancer preventative services), Comprehensive post-abortion care (CAP), labor and delivery family planning and Prevention of mother-to-child transmission of HIV (PMTCT). In cases where pregnant women and newborns face life-threatening complications—such as severe bleeding, infection, prolonged or obstructed labor, eclampsia, and newborn asphyxia—comprehensive emergency obstetric and newborn care services (CEmONC) are provided. Additionally, there are services available for family planning, PMTCT, Basic Emergency Obstetric and Newborn Care (BEMONC).

In addition, data were collected through document reviews, including electronic records with aggregated data and standardized paper-based Health Management Information Systems (HMIS) across all health facilities in the region. These data encompassed perinatal data recording systems, maternal healthcare services provided at each level, how data are utilized for clinical decision-making, and any relevant policy changes or reforms.

#### Registration of women at the Reproductive and Child Health Clinic (RCH) in the region

In 2021, a total of 49,380 women were registered at antenatal care services in the region. Nearly half, 22,980 (46.5%) had their first antenatal care (ANC) visit at less than 12 weeks (i.e. early ANC booking), more than half 26,400 (53.5%) had late ANC after 12 weeks gestation. (**Table 2**).

A total of 45343 births were recorded in the region, of these, 0.1 % (53/45343) were attended by traditional birth attendants. Only 0.7% (334/45343) of the births occurred before reaching the hospital, 0.2% (105/45274) occurred at home and most 98.9% (44782/45343) were facility deliveries. Among the facility deliveries, 2843 occurred at the zonal hospital and 2230 at the regional referral hospital (**Table 3**). Caesarean sections were performed in 20.3% (9186/45343) of births, primarily at the hospitals in Moshi Municipal District (**Table 4**). The neonatal death rate was 10.1 deaths per 1000 live births ((456/45274) *1000) and the maternal death rate was 146 deaths per 100,000 live births ((66/45274) *100000) (**Table 5**).

#### Trends of maternal, and neonatal mortality (2020-2023)

The maternal mortality rate was 112 deaths per 100,000 live births ((51/45507)*100000) in 2020, 146 deaths per 100000 live births ((66/45177)*100000) in 2021, and 75 deaths per 100000 live births ((39/51514)*100000) in 2022, and 51 deaths per 100000 live births (37/72129)*100000) in 2023 (**Figure 2**).

The neonatal mortality trend was 9.5 deaths per 1000 live births ((432/4507)*100000) in 2020, 10.1 per 1000 live births ((456/45177)*100000) in 2021, 10.1 per 1000 live births ((366/51514)*1000) in 2022, 3.7 per 1000 live births ((273/72129)*1000) in 2023 (**Figure 3**). (Data from the Kilimanjaro regional office date 13 June 2024).

#### Registration of women at the Reproductive and Child Health Clinic (RCH) in the region

In 2021, a total of 49,380 women were registered for antenatal care services in the region. Nearly half, 22,980 (46.5%) had their first antenatal care (ANC) visit at less than 12 weeks of gestation (i.e. early ANC booking), the remaining 26,400 (53.5%) had their ANC after 12 weeks of gestation, reflecting late ANC (**Table 2**).

A total of 45343 births were recorded in the region whereby, 0.1 % (53/45343) of deliveries were attended by traditional birth attendants, 0.7% (334/45343) occurred before reaching the hospital, 0.2% (105/45274) occurred at home and 98.9% (44782/45343) were facility deliveries. There were 2843 births at the zonal hospital and 2230 at the regional referral hospital (**Table 3**). There is a difference of 4037 (8.2%) and the number of women registered for ANC. This could be explained by the pregnancy loss or having birth outside the region.

Among the deliveries, 20.3% (9186/45343) were by cesarean section (**Table 4**). The neonatal death rate was 10 deaths per 1000 deliveries ((456/45274) *1000) and the maternal death rate was 145 deaths per 100,000 deliveries ((66/45274) *100000) (**Table 5**).

#### Findings of the Exploratory Qualitative Study

Five themes were identified which provided insights into practices of data collection and utilization at different levels of the Tanzanian health system. Table 6 summarizes those findings, which are briefly outlined below.

#### Data Collection and Recording

Data is collected verbally and through clinical measurement by a nurse-midwife (NM); the government-mandated approach to data recording involves the use of four, paper-based, MTUHA books. A second approach involves dual recording in both MTUHA books and the digital system -DHIS2; this operates at district, regional, and national hospital levels, designated hospitals, and some of the identified health centers.

> “Yes, we do use both systems where health workers extract data from the MTUHA books and insert them into the DHIS2 later.” (DRCHCO 03)

The third approach; digital-only data recording, is used only in some private hospitals that utilize a variety of digital systems. NMs present a strong understanding of data collection and recording processes. Regardless of the level of health facility, and its public or private status, the types of data collected and recorded are highly standardized.

> “There is no big difference in information collected except the way they record the information” (LO 02).

#### Data Storage and Flow

MTUHA books are stored by NMs in locked cabinets, typically in consulting rooms which are locked, with limited staff access. A monthly summary data report is generated by each health facility on health indicators including frequency of ante-natal visits, number of deliveries, maternal and infant mortalities, pre/Eclampsia and post-partum hemorrhage, postnatal care, and child health. Paper-based reports are generated from MTUHA books where only they are in use and sent to the DRCHCO who inserts the data into DHIS2.

> “At the end of every month, there is a report prepared that is submitted at my office as a DRCHCO, and then from there I can insert the data into the DHIS2 system”. (DRCHCO 06)

In the dual-recording approach, monthly reports are generated in DHIS2. The DRCHCO reviews all summary reports, verifies data accuracy and completeness, and then shares and reviews reports with the DMO. Once stored in the DHIS2, data from each facility is accessed by the DMO’s office, the Regional Medical Officer’s office, and the Ministry of Health.

#### Challenges of Paper Records

Recording data in MTUHA books is time-consuming for NMs, especially when working alone.

> “It’s hectic, you ought to fill in all the books on your own during the clinic day, sometimes you forget to fill in a certain field” (NM 02)

Data loss and difficulties with data retrieval were also reported, for example,

> “We face the challenge of loss of data because we store the patients’ information in files within cabinets, if a patient comes in the later visits, it becomes hard to trace back past information”. (LO 03)
>
> *“The books can be torn resulting in loss of information and difficulty in tracing past information”* (NM03).

#### Data Usage in Clinical Decision-Making

Nurse Midwives and Lead Obstetricians are the primary users of data for clinical decision-making concerning the well-being and management of pregnant women, healthy babies, and mothers’ post-partum. NMs manage typical pregnancies, labor, and deliveries while Lead Obstetricians manage more complex cases. Analysis of monthly summary data informs decision-making by DMOs and health facility Heads of Departments. For example:

> “….. We have analyzed the data and found that many mothers are late starting their first clinic visit under 12 weeks. Once you know they are late, you must start looking for the reason ……” (DRCHCO 04)

#### Data Usage in Policy and Guideline Development

Informed by trends in health indicator data, DMOs reported how they make recommendations for changes in policies and guidelines at regional level to the Regional Medical Officer, and at national level to the Ministry of Health, and the ministry known as the ‘President’s Office, Regional Administration, and Local Government’ (PORALG), per the Tanzania Health Sector Strategic Plan July 2021-June 2026 (known as HSSP V). Final decisions regarding policy or guideline amendments or implementation are a function of the ministries.

## Discussion

We conducted this study to describe the distribution of births and adverse pregnancy outcomes, the maternal and newborn health services, the data recording system, and the experiences of those who record and use the data for clinical and policy decision-making. The data from 2021, suggest that the rates of maternal mortality, neonatal mortality, and stillbirth were 146 per 100,000 births, 10 per 1000 live births, and 4 per 1000 births, respectively. The great majority of these deaths occurred in the Moshi Municipal, which reflects the fact the Kilimanjaro Clinical Medical Centre (KCMC) zonal and regional referral hospitals are in this district. Since the maternal mortality rate and neonatal mortality rate in 2021 was high to gain thorough understanding, data from 2020 and also for 2022 and 2023 was reviewed. Both data indicated a similar pattern where there is first increase in mortality rates from 2020 to 2021, then by a sharp decrease in 2022 and addition decrease in 2023.

The Cesarean section rate is 20%, which seems to be distributed relatively equally across the districts ranging from 14.8% in Siha District and 19.5% in Rombo District. However, the Cesarean section rate is highest in Moshi Municipal District at 29.3%, which is not surprising considering that the KCMC zonal referral hospital, which services the whole of North Tanzania, and the Kilimanjaro Regional Hospital are in this District. The low uptake of early ANC remains a concern with half of the pregnant women starting their ANC after 12 weeks’ gestation.

Maternal mortality ratio varies across countries in Sub-Sahara Africa (Alebel *et al*., 2020). The study found the maternal mortality was 146 per 100,000 which is higher than the maternal mortality national estimate, using the Tanzania Demographic Health Survey, of 104 deaths per 100,000 (95% CI: 59, 149). The survey uses a two-stage sample design, in the first stage enumeration areas from the 2012 census were selected to obtain 629 clusters from both urban and rural. In the second stage, 26 households were selected systematically from each cluster to obtain the required sample size (Ministry of Health (MoH)[Tanzania Mainland] *et al*., 2022). The difference can be explained by the nature of the studies, this study was a cross-sectional hospital-based study.

The regional maternal mortality rate of 146 deaths per 100,000 is lower than the estimate from the global maternal report of 238 per 100,000 (WHO, 2021). The possible explanation could be due to methodological differences in the data collection of the two estimates. The estimated rate from this study is based on the hospital data with the possibility of underestimation of unreported deaths such as death at home. But also, this could be due to the validity, incompleteness, and accuracy of the data according to our qualitative study findings. The global maternal estimates have multiple sources which are civil registration and vital statistics, specialized studies on maternal mortality such as confidential inquiries into maternal deaths, and reproductive-age mortality studies. Also, surveys and other miscellaneous data sources are used with approval from country-level (WHO, 2021). This highlights the difficulties of estimating maternal mortality without robust digital health information systems. The maternal mortality rate is higher than the target rate WHO target of 72 deaths per 100,000 births (United Nations: Department of Economic and Social Affairs, 2022). The variations can be due to geographical locations and the political efforts done by the country to reduce the maternal mortality rate.

The reported neonatal mortality rate in this study of 10 per 1000 which is lower compared to the TDHS report of 24 deaths per 1000 live births in a country. The difference can be due to underreporting of the cases, as TDHS data is community-based hence deaths were more likely to be captured than in this study based on paper-based healthcare facility records. It is also lower than the global neonatal mortality ratio of 17.9 deaths per 1000 births (Sharrow *et al*., 2022), the variation could be due to the different estimation procedures. The global estimate was based on subnational level data from 1990 to 2019 inclusive (GBD, 2019) and the country data are healthcare facility-based which are mostly early neonatal deaths. Similar to the maternal mortality rate, it is possible that neonatal deaths at home are not captured in the regional health information system.

The trend in maternal and neonatal mortality can be explained by the following the first increase from 2021 due to COVID-19 Pandemic, this could be due increased complications during childbirth due to the pandemic. And also healthcare staff shortages could have caused the higher mortality rates. And the decrease from 2021 to 2023 could be due to improved healthcare services, widespread of COVID-19 vaccines and target interventions in the region such as M-mama. Which target to facilitate transportation for pregnant women to prevent their delay to care (Njiro *et al*., 2023; Munishi *et al*., 2023).

The reported stillbirth in this study of 4 deaths per 1000 births is lower compared to that reported in the Tanzania Demographic Health Survey of 37 deaths per 1000 births (Ministry of Health (MoH)[Tanzania Mainland] *et al*., 2022). This can be explained by the difference in study designs as above.

The cesarean section rate of 20.3% in the region is high compared to the recommended rate of between 10% and 15% by the WHO (The Lancet, 2018). Most of the cesarean sections are done at the referral hospitals which mostly receive complicated cases. This difference may be explained by overutilization of the service which may put the mothers and newborns at unnecessary risk such as surgical site infection, intra-operative bleeding, anemia, and post-partum hemorrhage. But also this poses a need for policymakers to review the policies and provide education to reduce the rate of cesarean section to preserve resources and prevent adverse effects on mothers and newborns (Dibabi *et al*., 2022). In addition, the data which was collected used MTUHA books which might lead to over or under estimations of the true challenge and this pose the need to have Electronic Health Record System (EHRS).

ANC is an important component in the reduction of maternal mortality. The WHO recommends a minimum of four ANC visits during pregnancy. The focus of antenatal care in the country is to ensure pregnant women attend their first ANC visit within the first 12 weeks of pregnancy (WHO, 2016; MoHCDGEC, 2018). The visit should be within the required and proper timing, the services are from qualified healthcare providers. Also, pregnant women should receive the services such as blood pressure measurement, urine sample, blood sample, listening for the baby’s heartbeat, counseling about the mother’s diet, counseling about breastfeeding, and checking for vaginal bleeding (MoHCDGEC, 2018).

Clinical decision-making is central to improvements in clinical practice and care for pregnant women and newly born children. Qualitative findings indicate that in clinical decision-making by nurse midwives the primary use data is in caring for the individual pregnant woman, with the aim of safe deliver of a healthy baby and healthy mother. For major complications, such as excessive bleeding, they do not have the authority to make clinical decisions and must consult a medical officer. Data is used in clinical practice by lead obstetricians also, to manage normal, and complicated pregnancies, labor, and deliveries.

District medical officers (DMOs) make budgetary decisions and policy recommendations based on the health indicators such as number of deliveries, maternal mortalities, child mortalities, common causes of major complications like pre/eclampsia, post-partum hemorrhage.

In conclusion, the Kilimanjaro region has a high burden of maternal and neonatal mortality but less in comparison with national target, which is likely to be under-reported by the current national health information system. The increasing caesarean section rate is an important development which will likely improve maternal and neonatal outcomes, but it needs to examine to ensure health equity. On the other hand, the low uptake of early ANC is concerning and needs to be improved. Paper-based data collection faces limitations such as manual entry errors, difficulty in accessing large volumes, physical storage demands, vulnerability to damage or loss, labor-intensive integration into digital systems, and potential delays in updating information. These constraints highlight the inefficiencies and risks associated with relying solely on paper-based methods for modern contexts that prioritize accuracy, accessibility, and timeliness of information.

## Data Availability

Researchers can apply to access data from the Regional Medical Office of Kilimanjaro region by contacting the Regional Administrative Secretary in Tanzania, followed by submission of a proposal. The data are not publicly available due to privacy/ethical restrictions and only available upon reasonable request.

## Author’s contributions

ASK, BTM, SW and CR designed the study. MM, FMP, MC and DB drafted the manuscript. GAO, GN, AS, FR, GN, MM, and FMP did quantitative data gathering. ASK, MM and MJM performed the statistical analysis for quantitative part. FMP, LM and JS participated in qualitative interviews, transcription, and translation. FMP, LM, SW and MC did the qualitative data synthesis. All co-authors participated in editing the manuscript and approved the final manuscript for submission.

## Acknowledgement

We thank the participants of the qualitative study for their time and contributions.

## Appendix

### Tables and figures

**Table 1:**
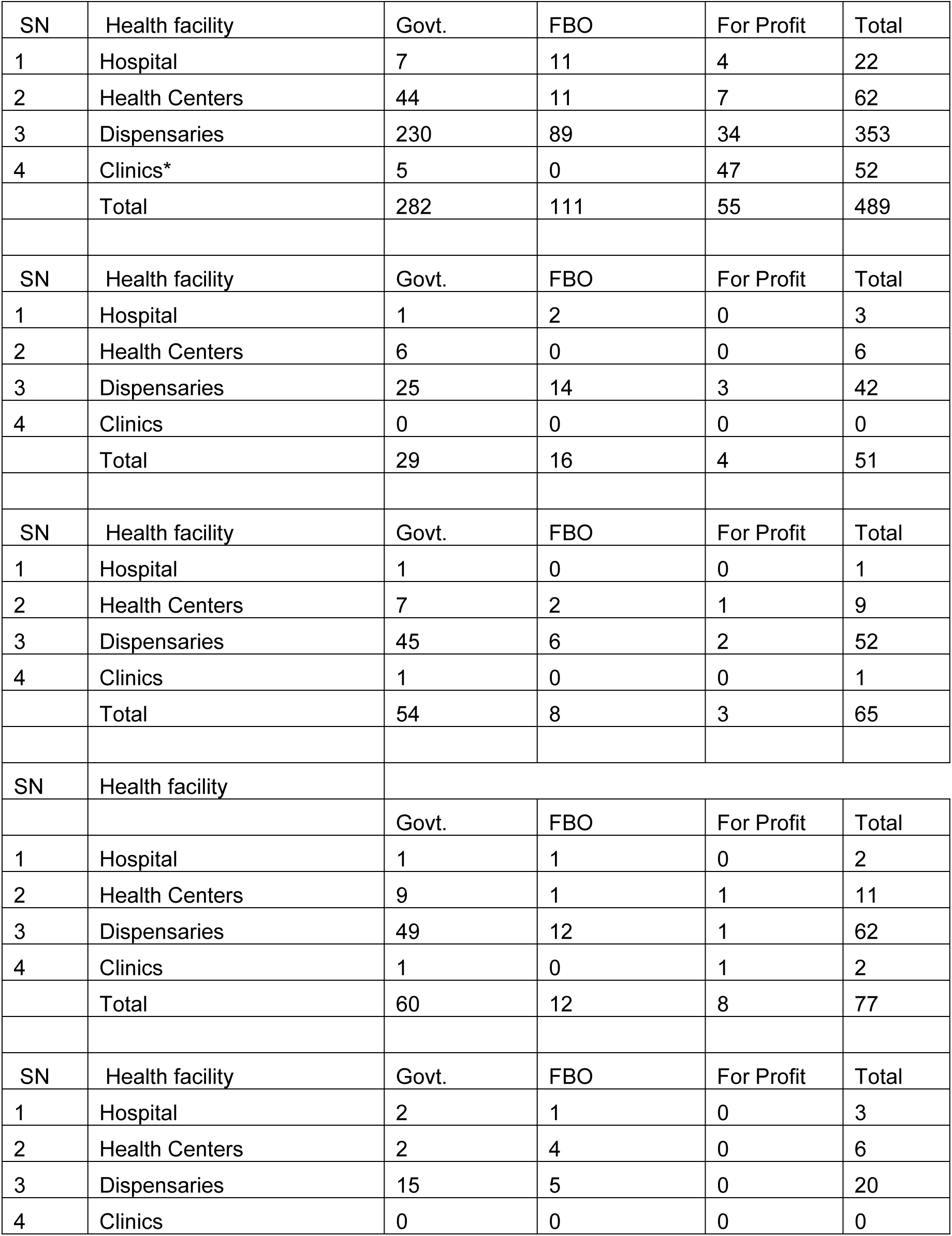
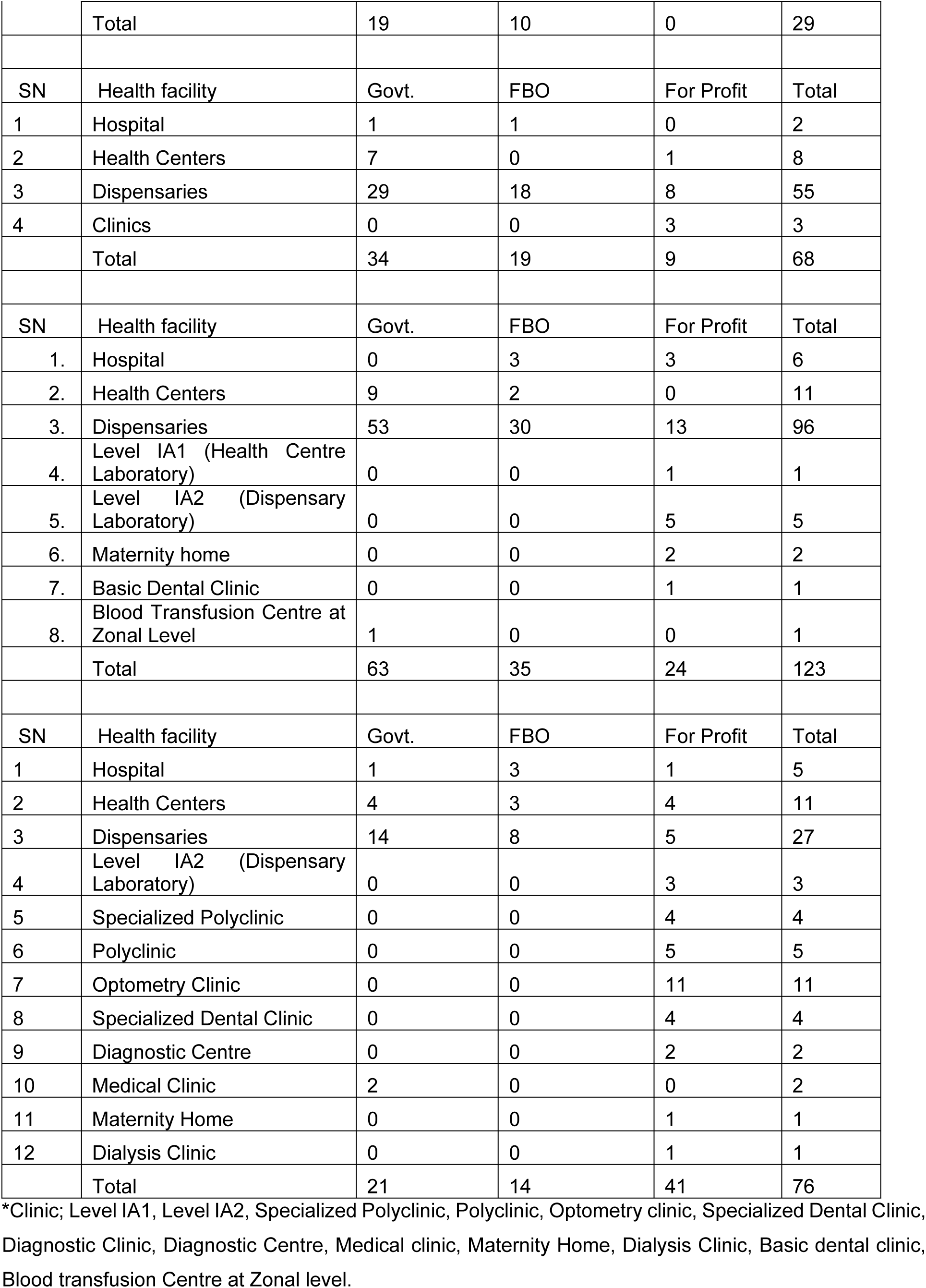
Regional health facilities by district.

**Table 2:** Women attendance of first ANC visit in Kilimanjaro region (N=49380).

| **Area** | **1<sup>st</sup> ANC Visit**<br>**<12 GA weeks n (%)** | **1<sup>st</sup> ANC Visit**<br>**≥ 12 GA week n (%)** | **1<sup>st</sup> ANC Visit**<br>**(total)** |
| --- | --- | --- | --- |
| Hai District | 1445(22.7) | 4929(77.3) | 6374 |
| Moshi DC (Moshi Rural) | 5302(46.7) | 6040(53.3) | 11342 |
| Moshi MC (Moshi Urban) | 4273(42.9) | 5689(57.1) | 9962 |
| Mwanga District | 1519(43.2) | 1994(56.8) | 3513 |
| Rombo District | 3359(53.2) | 2950(46.8) | 6309 |
| Same District | 5736(72.4) | 2189(27.6) | 7925 |
| Siha District | 1346(34.0) | 2609(66.0) | 3955 |
| Kilimanjaro Region | 22980(46.5) | 26400(53.5) | 49380 |

**Table 3:**
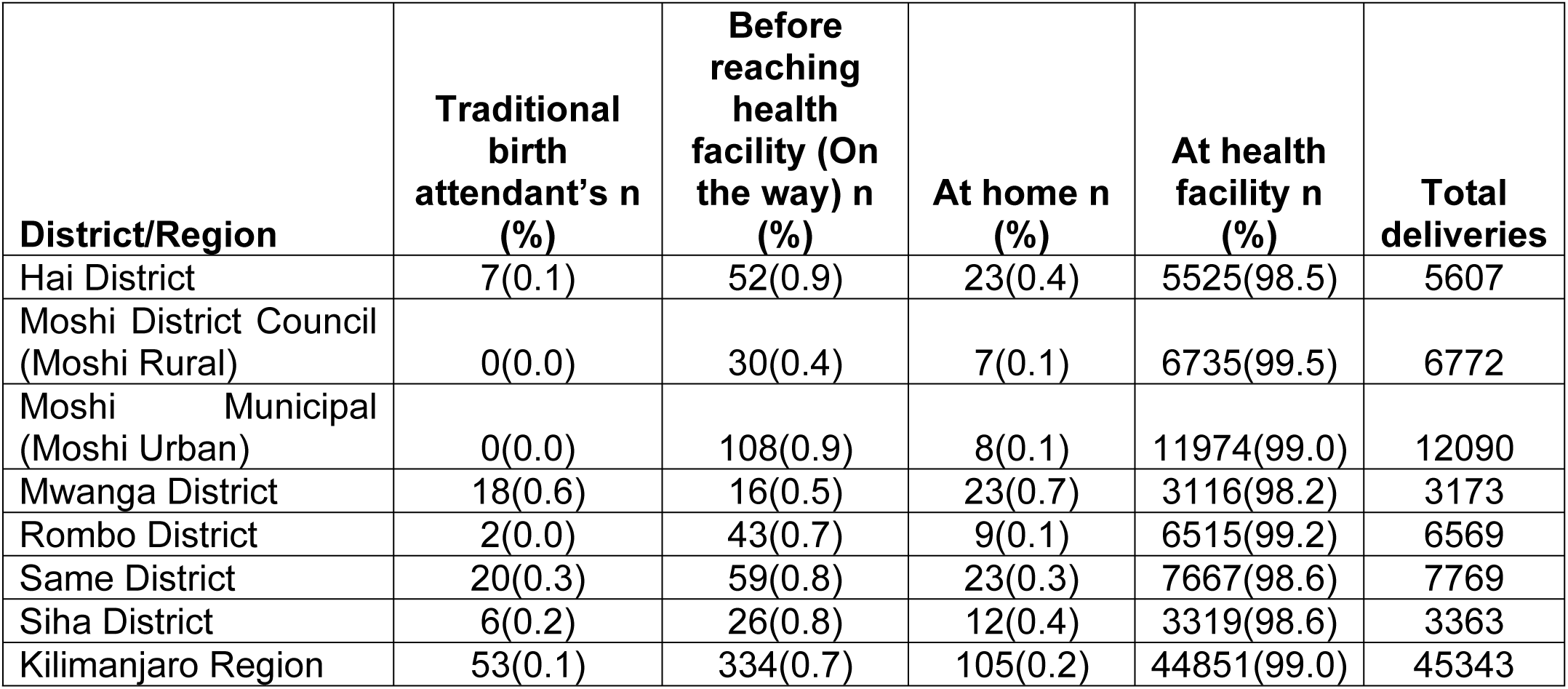
Number of deliveries in the Kilimanjaro region, by place of delivery (N=45343)

| **District/Region** | **Traditional**<br>**birth**<br>**attendant's n**<br>**(%)** | **Before**<br>**reaching**<br>**health**<br>**facility (On**<br>**the way) n**<br>**(%)** | **At home n**<br>**(%)** | **At health**<br>**facility n**<br>**(%)** | **Total**<br>**deliveries** |
| --- | --- | --- | --- | --- | --- |
| Hai District | 7(0.1) | 52(0.9) | 23(0.4) | 5525(98.5) | 5607 |
| Moshi District Council<br>(Moshi Rural) | 0(0.0) | 30(0.4) | 7(0.1) | 6735(99.5) | 6772 |
| Moshi Municipal<br>(Moshi Urban) | 0(0.0) | 108(0.9) | 8(0.1) | 11974(99.0) | 12090 |
| Mwanga District | 18(0.6) | 16(0.5) | 23(0.7) | 3116(98.2) | 3173 |
| Rombo District | 2(0.0) | 43(0.7) | 9(0.1) | 6515(99.2) | 6569 |
| Same District | 20(0.3) | 59(0.8) | 23(0.3) | 7667(98.6) | 7769 |
| Siha District | 6(0.2) | 26(0.8) | 12(0.4) | 3319(98.6) | 3363 |
| Kilimanjaro Region | 53(0.1) | 334(0.7) | 105(0.2) | 44851(99.0) | 45343 |

**Table 4:**
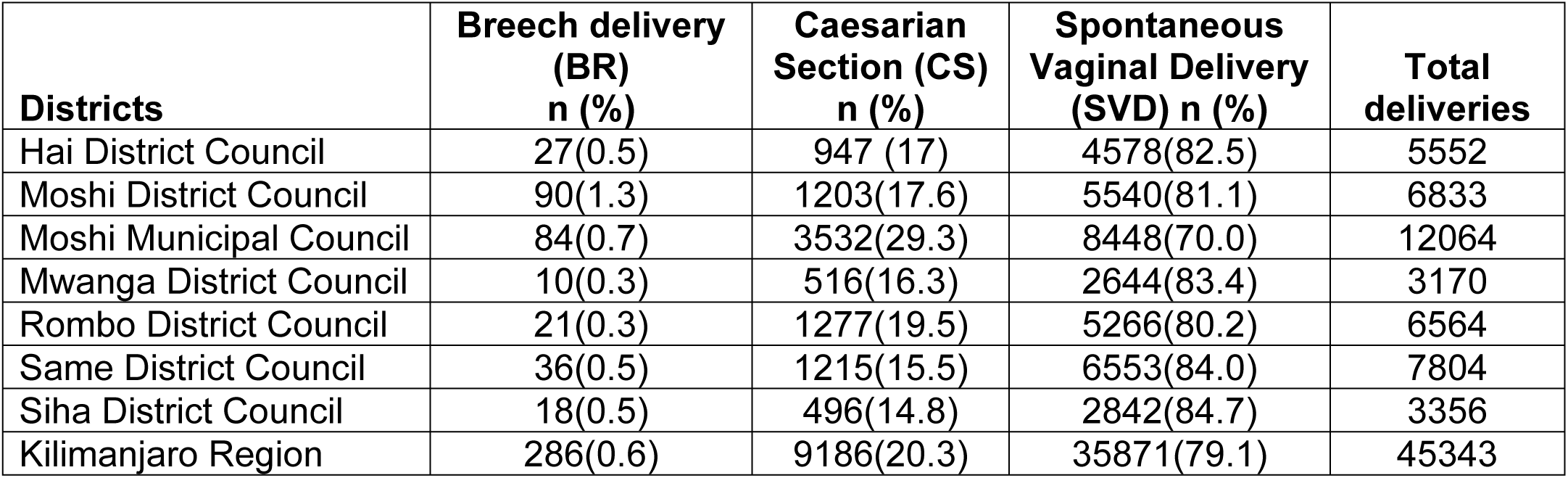
Number of deliveries in the Kilimanjaro region, by mode of delivery (N=45343)

| **Districts** | **Breech delivery**<br>**(BR)**<br>**n (%)** | **Caesarian**<br>**Section (CS)**<br>**n (%)** | **Spontaneous**<br>**Vaginal Delivery**<br>**(SVD) n (%)** | **Total**<br>**deliveries** |
| --- | --- | --- | --- | --- |
| Hai District Council | 27(0.5) | 947 (17) | 4578(82.5) | 5552 |
| Moshi District Council | 90(1.3) | 1203(17.6) | 5540(81.1) | 6833 |
| Moshi Municipal Council | 84(0.7) | 3532(29.3) | 8448(70.0) | 12064 |
| Mwanga District Council | 10(0.3) | 516(16.3) | 2644(83.4) | 3170 |
| Rombo District Council | 21(0.3) | 1277(19.5) | 5266(80.2) | 6564 |
| Same District Council | 36(0.5) | 1215(15.5) | 6553(84.0) | 7804 |
| Siha District Council | 18(0.5) | 496(14.8) | 2842(84.7) | 3356 |
| Kilimanjaro Region | 286(0.6) | 9186(20.3) | 35871(79.1) | 45343 |

**TABLE 5:** Maternal and child delivery outcomes in Kilimanjaro region, 2021 (N=45343)

| **Maternal**<br>**and child**<br>**delivery**<br>**outcomes** | **Macerated**<br>**still birth**<br>**(Per 1000**<br>**births)** | **Fresh still**<br>**birth (Per**<br>**1000 births)** | **Still birth n**<br>**(Per 1000**<br>**births)** | **Neonatal**<br>**death (Per**<br>**1000 live**<br>**births)** | **Maternal**<br>**death (Per**<br>**100,000**<br>**births)**<br>**(95%CI)** |
| --- | --- | --- | --- | --- | --- |
| **Moshi**<br>**district** | 0.1(0.0,0.8) | 0.1(0.0,0.8) | 0.3(0.0,1) | 0.1(0.0,0.8) | 14.6(0.37,81.5) |
| **Moshi**<br>**Municipal** | 6.6(5.2,8.2) | 4.4(3.3,5.8) | 11.1(9.3,13.1) | 34.6(31.4,38) | 460(347,590) |
| **Hai district** | 0.5(0.1,1.6) | 0.7(0.2,1.8) | 1.3(0.5,2) | 2.5(1.4,4.2) | 0(0,0) |
| **Siha district** | 0.3(0.0, 1.7) | 2.4(1.0,4.6) | 2.7(1.2,5) | 2.1(0.8,4.3) | 59.7(7.2, 210) |
| **Rombo**<br>**district** | 1.1(0.4,2.2) | 0.5(0.1,1.3) | 1.5(0.7,2.7) | 0.6(0.2,3.2) | 61(16.6,150) |
| **Mwanga**<br>**district** | 0.9(0.2,2.8) | 0.3(0.0,1.7) | 1.3(0.3,3.2) | 1.3(0.1,2.2) | 63.2(7.7,220) |
| **Same**<br>**district** | 0.1(0.0,0.7) | 0.1(0.0,0.7) | 0.3(0.03,0.9) | 1(0.4, 2) | 25.6(3.1,90) |
| **Total** | 2.1(1.7,2.5) | 1.6(1.2,1.9) | 3.7(3.1,4.2) | 10(9.2, 11) | 146(113,180) |

**\*The denominator for still births was births and denominator for maternal and neonatal deaths are live-births**

**Table 6:** Maternal and neonatal outcomes in Kilimanjaro region from 2020 to 2023.

| **Maternal and child delivery outcomes** | **Number of live birth/Total number of live births** | **Neonatal death (Per 1000 live births)** | **Maternal death (Per 100,000 births)** |
| --- | --- | --- | --- |
| 2020 | 45507 | 9.5(8.6, 10.4) | 112(83.5, 147.3) |
| 2021 | 45177 | 10.1(9.2,11) | 146(113,180) |
| 2022 | 51514 | 7.1(6.4,7.9) | 75.7(53.8,103.4) |
| 2023 | 72129 | 3.7(3.3,4.3) | 51.3(36.1, 70) |

**Table 7:**
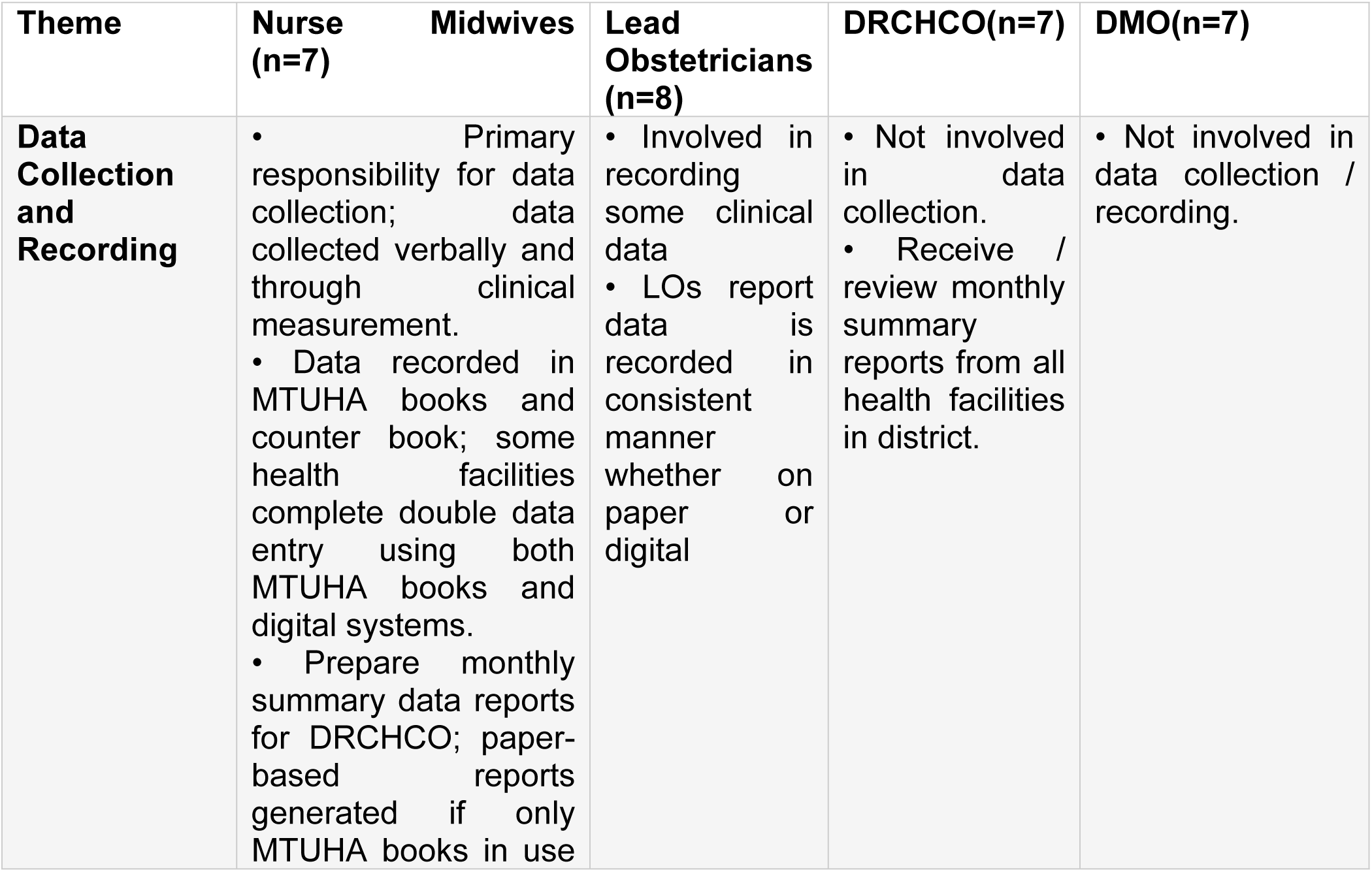
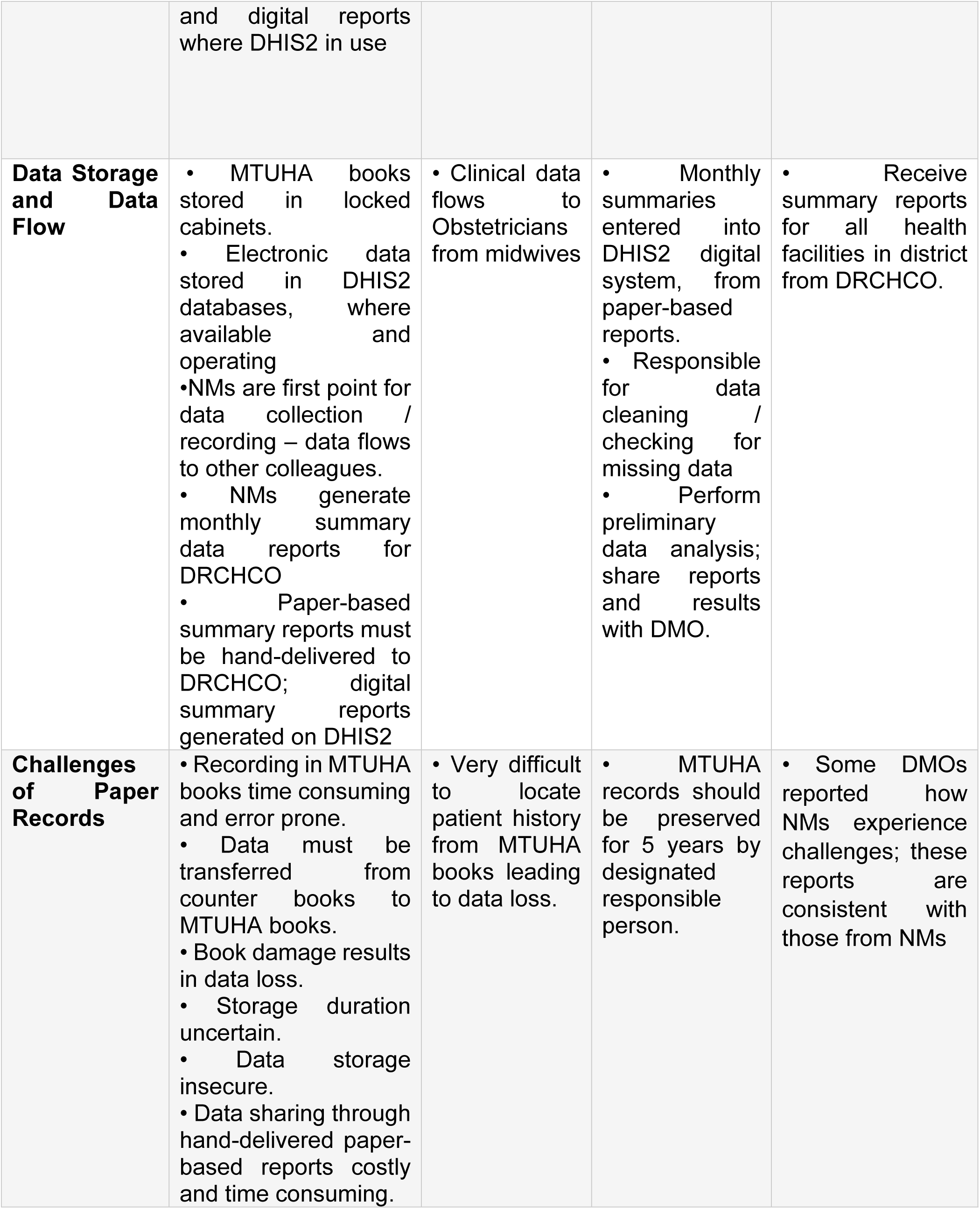
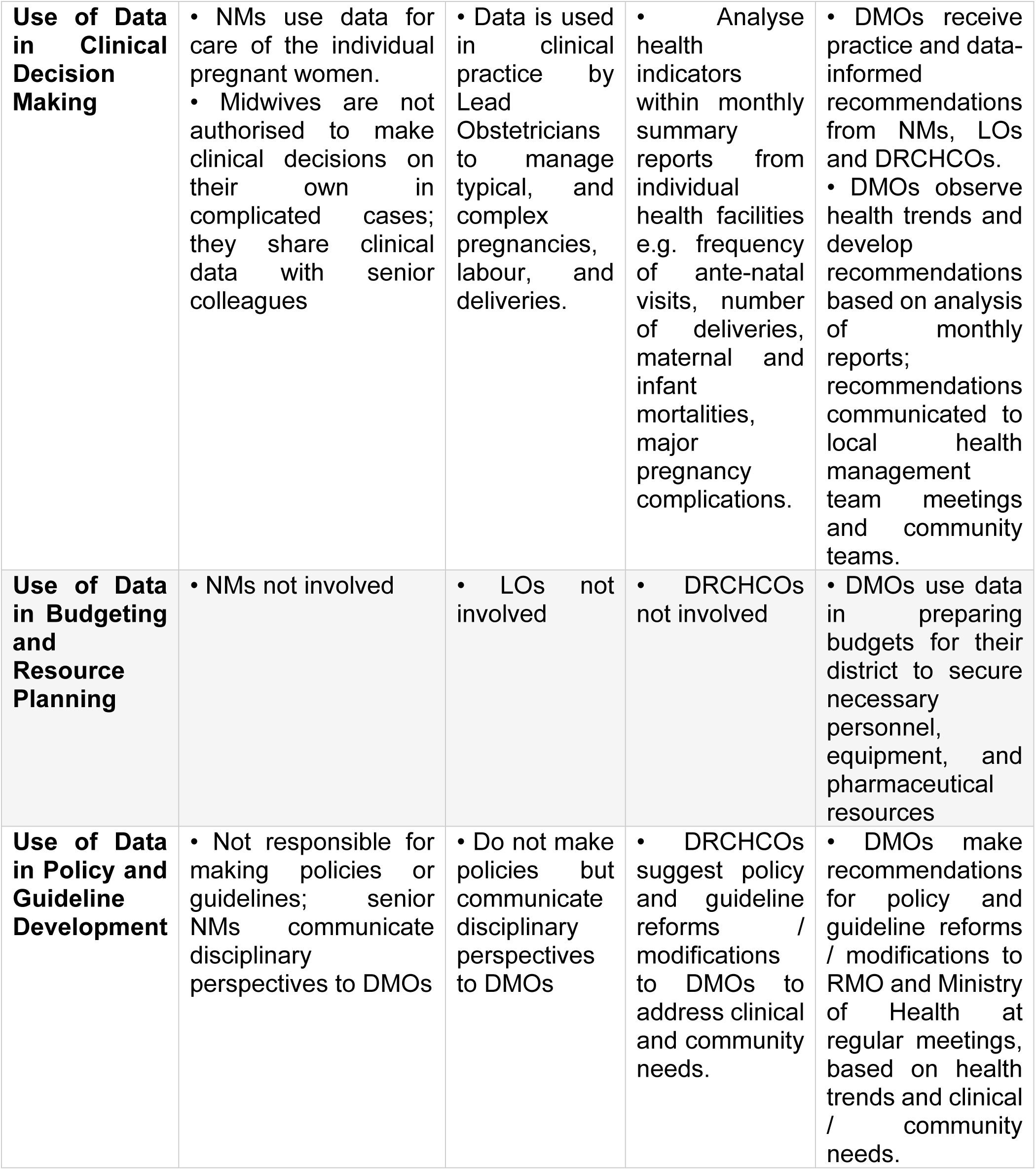
Summary of Qualitative Findings, organized by Theme and Interview Group.

**Figure 1:**
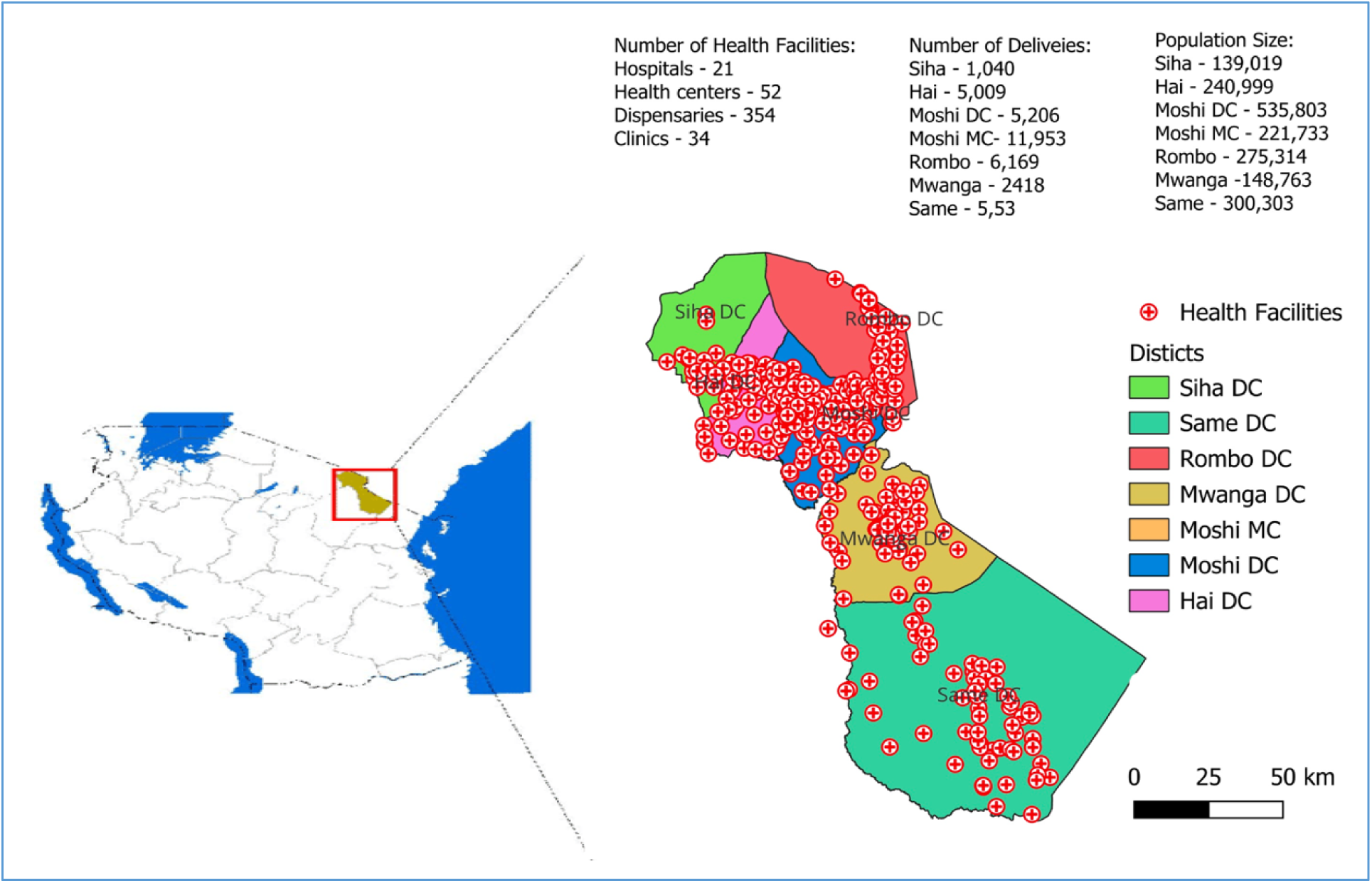
Heat map showing the distribution of health facilities in the Kilimanjaro region.

**Figure 2:**
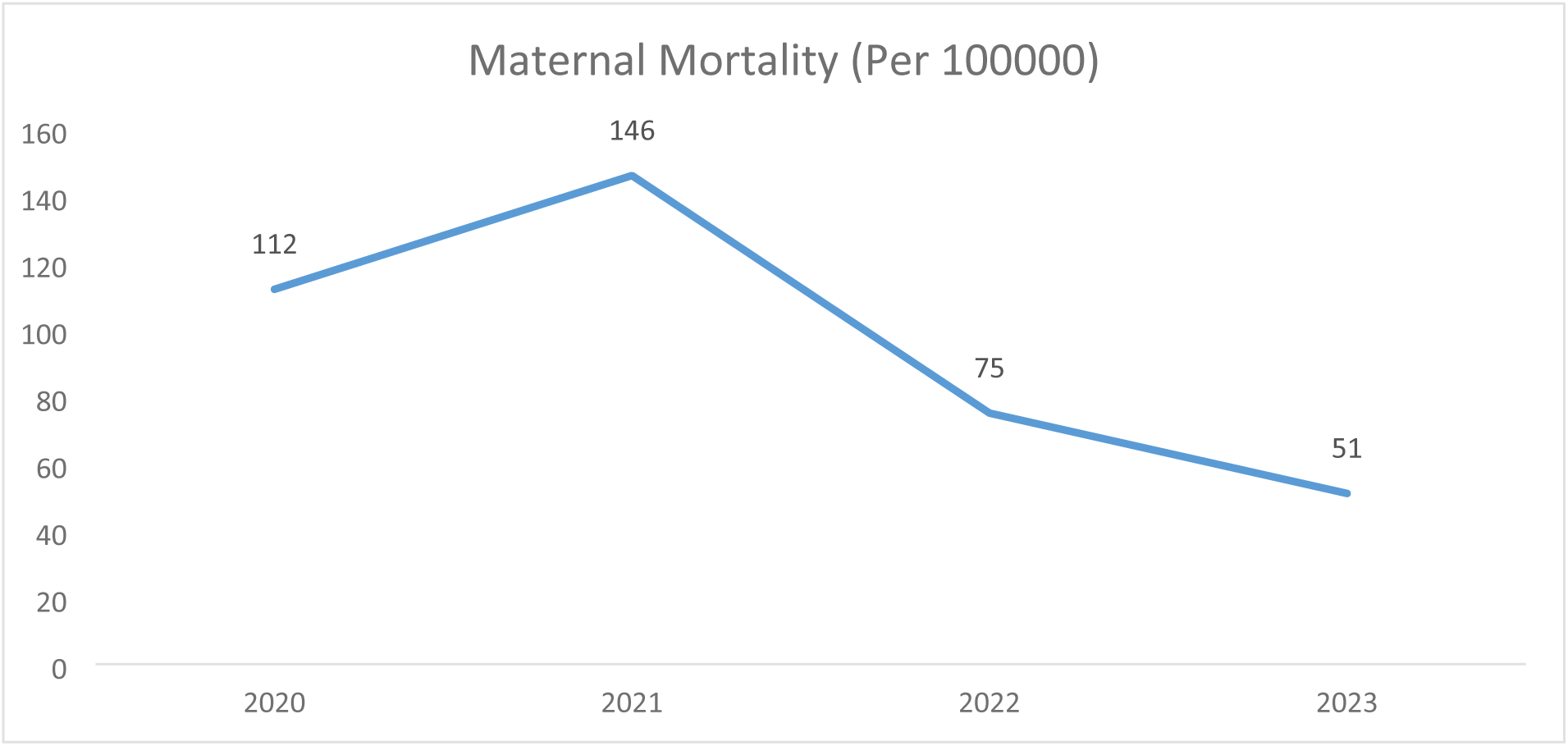
The trend of maternal mortality in Kilimanjaro region from 2020 to 2023.

**Figure 3:**
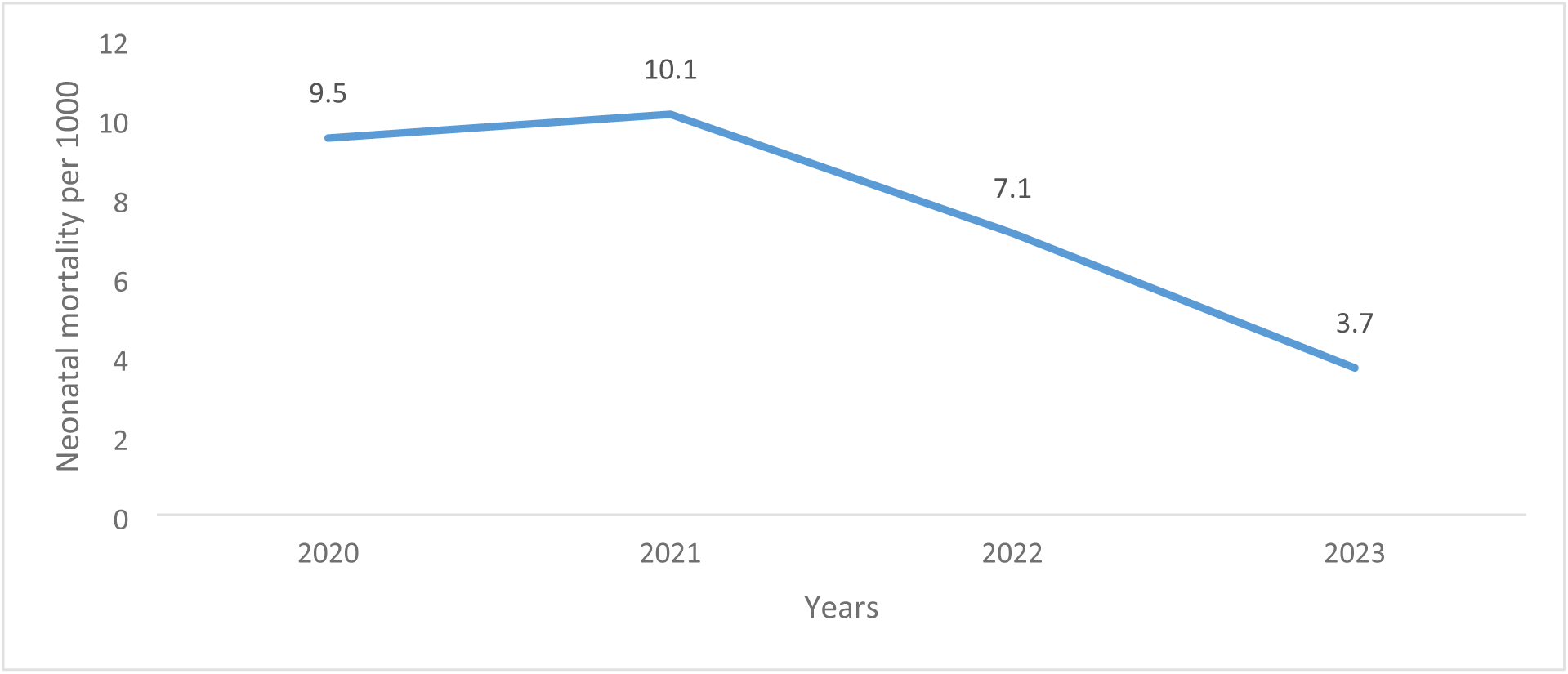
The trend of neonatal mortality in Kilimanjaro region from 2020 to 2023.

